# A study on the Component Isochronic substitution Benefits of 24-hour Activity Behavior on Emotional and Behavioral Problems in Preschool Children

**DOI:** 10.1101/2025.11.24.25340914

**Authors:** Qiong Shen, Aolin Yu

**Author notes:** Corresponding author: Aolin Yu.

## Abstract

**Objective:** To explore the association between 24-hour activity behavior and emotional behavior problems using the component data isochronic substitution model, and to analyze the expected changes in emotional behavior problems of preschool children after different activities are substituted for each other.

**Method:** The physical activity, sedentary and sleep data of 211 preschool children aged 3 to 6 (91 boys and 120 girls) were measured using the ActiGraph wGT3-BT accelerometer, and the emotional and behavioral problems of preschool children were evaluated using the Strengths and Difficulties Questionnaire (SDQ). Descriptive statistical analysis of component data and isochronous substitution were both accomplished using R software.

**Result:** 1) The MVPA time of preschool children is seriously insufficient. There are significant differences in MVPA between different genders (P < 0. 01). The MVPA time of boys is significantly higher than that of girls. There are significant differences in MVPA (P < 0. 01) and SP (P < 0. 05) among different ages. 2) The abnormal detection rate of the total score of emotional and behavioral problems and difficulties among preschool children was 12. 8%, while the abnormal detection rates of emotional symptoms, conduct problems, hyperactivity and inattentiveness, peer interaction problems, and prosocial behaviors were 13. 8%, 13. 8%, 17. 7%, 21. 4%, and 5. 3% respectively. 3) The simultaneous replacement of LPA, SP, SB at 15 minutes of mVPA and the replacement of SP with LPA would alleviate the emotional and behavioral problems of preschool children (P < 0. 05), while the opposite would aggravate the emotional and behavioral problems of preschool children (P < 0. 05). 4) The substitution results of different behaviors are asymmetrical, that is, emotional and behavioral problems are prone to occur but difficult to alleviate.

**Conclusion:** The distribution of 24-hour activity and behavior time is closely related to the emotional and behavioral problems of preschool children. Although MVPA has the greatest effect on alleviating the emotional and behavioral problems of preschool children, it is more necessary to rationally allocate the influence of the use of each activity and behavior time on the emotional and behavioral problems of preschool children from the overall perspective of 24-hour activity and behavior. In the environments of kindergartens, families and communities, it is emphasized to transform children’s SB and LPA into MVPA, so as to better improve their emotional and behavioral problems.

## Introduction

Emotional and behavioral problems refer to abnormal behaviors that are beyond the normal range allowed by the age of the individual in terms of severity and duration [1]. Preschool children are in a sensitive period of physical and mental development [2–3]. During this period, the physical and mental functions of young children have not yet matured and are prone to emotional and behavioral problems due to the influence of many factors such as biology, family environment and society [4–5]. Emotional and behavioral problems not only have a negative impact on the social interaction ability and academic performance of preschool children themselves, but also cause adverse effects on the entire family and even society. Longitudinal evidence has found that preschool children with emotional and behavioral problems are associated with more severe psychological problems during school age and adolescence [6]. In recent years, growing evidence has shown that Physical Activity (PA), Sleep Period (SP), and Sedentary Behavior (SB) play unique roles in emotional and behavioral problems in preschool children [7–9]. However, these studies explored the influence of a single activity behavior on the emotional and behavioral problems of preschool children from a “fragmented” perspective, ignoring the intrinsic connections among various activity behaviors [10]. The isochronic substitution model of component data adopts Isometric log-ratio (ILR) to overcome the summation constraints of 24-hour activity behavior time data. On this basis, it simulates the mutual substitution of pairwise activity behaviors at the same time. It provides a more scientific and reasonable method for exploring the proportion of 24-hour activity behavior time and the impact of redistribution on health indicators from an overall perspective [11]. Although, at present, studies have explored the association between the attainment of 24-hour activity behavior and emotional and behavioral problems in different ages and different populations, few have adopted the component data isochronic substitution method to analyze the impact of the time allocation of 24-hour activity behavior on emotional and behavioral problems [12, 13]. Research on the 24-hour activity behavior and emotional behavior problems of preschool children is even more scarce.

Based on this, this study adopts the component data isochronic substitution method to objectively measure the 24-hour activity and behavior time of preschool children. Through the component isochronic substitution model, it explores the expected changes in emotional and behavioral problems of preschool children after mutual substitution among different behaviors. In order to provide theoretical support and practical guidance for improving the usage plan of 24-hour activity and behavior time and emotional and behavioral problems of preschool children in China.

## Research objects and methods

### Research Object

This study adopted the convenient sampling method and randomly selected 5 kindergartens in Zhuzhou area for sampling and investigation. This study followed the principle of voluntary participation. Before the investigation was carried out, the members of the research group issued informed consent forms to the guardians, informing them in detail of the research purpose, significance, test contents, procedures and relevant precautions, and asked the guardians to sign the informed consent forms. During the research process, guardians can withdraw at any time without reason. Inclusion criteria for subjects: 1) Age range: 3 to 6 years old;2) Good health, capable of participating in various activities, and no major growth and development diseases have been detected. 3) The guardian agrees to participate in this investigation. The missing samples of the 24-hour activity behavior monitoring data were eliminated, and the missing samples of the emotional behavior problem data were supplemented and improved in the later stage. A total of 300 preschool children aged 3 to 6 participated in this study. Among them, the 24-hour activity behavior monitoring data of 206 preschool children were valid data, and the emotional and behavioral problem questionnaires of 218 preschool children were valid data. Eventually, there were 206 valid data, with an effective rate of 68. 7%. The present study protocol was approved by the Ethics Review Committee of Wuxi Taihu University (Approval No: WXTC-IRB-2024-013). Written informed consent was obtained from a parent or legal guardian of each minor participant prior to the study.

## Research Methods

### Field measurement method

The recruitment of subjects for this study was conducted from March 1st to March 19th, 2024. A total of 300 preschool children were recruited to participate in this study, among which 206 valid data were obtained. The SB and Light physical activity of the research subjects were measured using the Actigraph wGT3X-BT Accelerometer. Levels of LPA, Moderate physical activity (MPA), and Vigorous physical activity (MPA) The time data of Moderate-to-vigorous physical activity (MVPA) is the sum of MPA and VPA time. This instrument has been proven to be an effective tool for objectively measuring the PA level of preschool children. After obtaining the consent of the subject and their guardian, the subject is required to wear the accelerometer continuously for one week (including five working days and two weekend days). After setting the sampling frequency, it is uniformly worn on the right anterior superior iliac spine. It is put on after getting up every day and taken off when going to bed at night. Except for taking a bath and doing water sports such as swimming, the instrument can be temporarily removed at all other times. During the 24-hour activity behavior monitoring process, the subjects maintained a normal living state. Before the measurement began, the researchers provided unified instrument guidance to the guardians and the kindergarten teachers accompanying the classes to ensure the accurate wearing of the instruments. The researchers retrieved the instruments on the 8th day after wearing them, and then used the ActiLife (Version 6. 13. 3) software for equipment initialization, data export and analysis. The specific parameter Settings are shown in Table 2. Sleep data preliminary investigation in the form of questionnaire, by parents to fill in four issues, including “children, working days at noon sleep time ___, ___ to ___, ___”, “children, weekday night sleep time ___: ___ to ___, ___”, “kids, the weekend at noon sleep time ___, ___ to ___, ___”, “children, Sleep time on weekend nights : ___ : ___ to ___ : ___. Finally, the sleep onset and wake-up time nodes are manually divided based on the data exported from ActiLife, and the sleep time is calculated accordingly.

**Table 1 Physical activity measurement parameter settings list of ActiGraph GT3X-BT**

### Questionnaire Survey Method

The Strength And Difficulty Questionnaire (SDQ) was adopted to assess children’s emotional and behavioral problems. This questionnaire has been widely used at home and abroad and requires parents to fill it out based on the behavioral performance of preschool children in the past six months [16]. The main part of the questionnaire consists of 25 scoring items. Each item is scored as 0, 1, and 2 respectively for non-conformity, partial conformity, and complete conformity. Among them, items 7, 11, 14, 21, and 25 are scored in reverse. The SDQ consists of a total of 5 factors, and the score range of each factor is from 0 to 10 points. These five factors are respectively emotional symptoms (including physical discomfort, tension in new environments, etc.), conduct problems (including temper tantrums, quarrels or bullying other children, etc.), hyperactivity and inattentiveness (including excessive activity, inattention, etc.), peer interaction, and prosocial behaviors (including consideration of others, willingness to help others, etc.) Among them, emotional symptoms and peer interaction problems belong to internalized behavioral problems, while conduct problems and hyperactivity factors belong to externalized behavioral problems. The sum of the scores of internalized behavior and externalized behavioral problems is the total difficulty score of SDQ. The higher the score, the more severe the existing objective difficulty degree. The higher the score of prosocial behavior, the better the prosocial ability.

According to Du [17] et al., the cut-off values of normal, marginal and abnormal for various factors of emotional and behavioral problems in preschool children aged 3 to 6 are as follows:The normal range of emotional symptoms is 0 to 3 points, 4 points is at the marginal level, and 5 to 10 points is abnormal. The normal range of conduct problems is 0 to 2 points, 3 points is at the marginal level, and 4 to 10 points is abnormal. Hyperactivity with abnormal attention is rated as 0 to 5 points, marginal as 6 points, and abnormal as 7 to 10 points. The normal score for peer interaction problems is 0 to 2 points, the marginal level is 3 points, and the abnormal score is 4 to 10 points. The normal prosocial behavior is 10 to 6 points, the marginal level is 5 points, and the abnormal level is 4 to 0 points. The normal total score of difficulty is 0 to 13 points, the marginal level is 14 to 16 points, and the abnormal score is 17 to 40 points.

### Statistical Analysis

The scores of emotional and behavioral problems of preschool children were descriptively analyzed using SPSS 27. 0 software. For the analysis of component data, this study followed the 24h activity component data analysis guidelines proposed by Chastin et al. [11] and used the Compositions package in R software (Version 4. 3. 0) to statistically analyze the component data. Firstly, the central tendency of the time usage components is described by calculating the geometric mean and arithmetic mean. Due to the characteristic of component data that a change in one type of data will inevitably lead to changes in other behaviors, the discrete trend must be reflected through the variance matrix (pairwise logarithmic ratio variance among different behaviors), thereby reflecting the dependence between different behaviors. The smaller the variance value, the higher the degree of dependence between the two; the larger the variance value, the lower the degree of dependence between the two. The composition characteristics of the 24-hour activity behavior of the test group are reflected by means of the ternary graph. Secondly, the time-equal substitution method of component data was adopted. Taking the 24-hour activity and behavior component data after ILR as the independent variable and the emotional and behavioral problems as the dependent variable, and controlling confounding factors such as gender and age, the predicted changes of emotional and behavioral problems in preschool children after the mutual substitution of ppairs of activity and behavior were explored. Previous studies have found that 15-minute isochronic substitution between ppairs of activity behaviors can have a significant impact on the corresponding outcome indicators [18]. Therefore, this study analyzed the predicted changes of emotional and behavioral problems in preschool children after 24-hour activity behavior and 15-minute isochronic substitution. Finally, for the models with significant differences in peer-to-peer substitution, a trend graph of change was drawn with an increment of 5 minutes to help explain the “dose-effect” relationship of the influence of mutual substitution between ppairs of activity behaviors on the emotional and behavioral problems of preschool children.

## Research Results and Analysis

### Descriptive Statistics

**Table 2 shows the central tendency and degree of dispersion of component data**

The measurement results of 24-hour activity behavior showed that the daily SP time of preschool children was 610. 1min, SB time was 438. 3min, LPA time was 356. 6min, and MVPA time was 35. 0min. The daily time proportions of SP, SB, LPA and MVPA were 42. 3%, 30. 4%, 24. 7% and 2. 4% respectively. There was no significant difference between the arithmetic mean proportion and the component mean proportion. From the variance matrix of the pairs logarithmic ratio of the 24-hour active behaviors, it can be known that the two behaviors closest to 0 in the matrix are highly proportional in terms of the time spent, and the higher the possibility of conversion at the same time, the pairs logarithmic ratio of SP and SB in the matrix of this study is the smallest, log (SB/SP) =0. 036, which means that the relationship between these two behaviors is the closest. The interdependence is the strongest, so SP and SB are most likely to switch in terms of time usage. Due to the existence of constants and constraints in the component data, a ternary graph (Figure 1) is adopted to describe the time distribution of the 24-hour activity behavior elements SP, SB, LPA, and MVPA of preschool children, representing the three behaviors in the form of a ternary graph matrix at one time.

**Figure 1.**
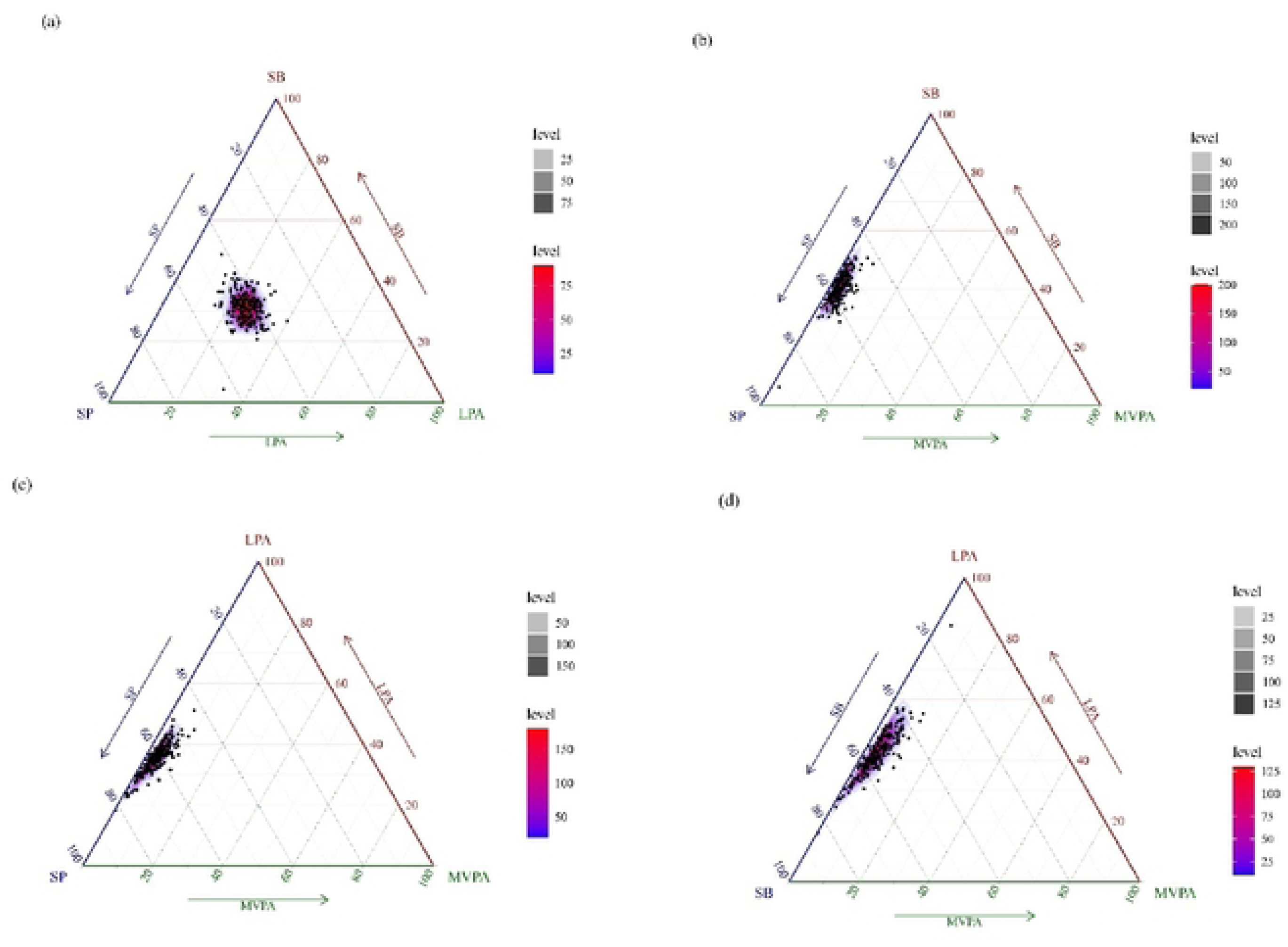
Ternary plots of sleep period (SP), sedentary behavior (SB), light physical activity (LPA) and moderate-to-vigorous physical activity (MVPA)

### Differences in 24-hour activity behaviors among preschool children of different genders and ages

Taking the time usage of SP, SB, LPA and MVPA as the dependent variables and gender as the independent variable respectively, one-way analysis of variance was used for the test. The results are shown in the table below. The differences in the daily MVPA time usage among preschool children of different genders were statistically significant (p < 0. 05). The average daily time spent by boys on MVPA was 40. 9 minutes, and that by girls was 30. 6 minutes.

**Table 3: Univariate analysis of 24-h movement behavior of school-age children of different genders**

Note: Bold font indicates statistical significance (P < 0. 05)

In terms of age structure, there was a statistically significant difference in the proportion of SP time among preschool children of different ages (p < 0. 05). There was also a statistically significant difference in the use of MVPA time among preschool children of different ages (p < 0. 01), and the MVPA time showed an upward trend with the increase of age.

**Table 4 Univariate analysis of 24-h movement behavior of school-age children**

Note: Bold font indicates statistical significance (P < 0. 05)

### Basic Information on Emotional and Behavioral Problems of Preschool Children

The results in Table 5 show that among the 207 preschool children surveyed, the average score of emotional symptoms was 2. 74±1. 78 points. Among them, the detection rates of normal, marginal and abnormal were 70. 9%, 17. 0% and 12. 1% respectively. The average score of peer interaction problems was 2. 43 ± 1. 57 points, and the proportions of normal, marginal and abnormal people were 55. 8%, 22. 8% and 21. 4% respectively. The average score of conduct problems was 2. 10± 1. 24 points, and the detection rates of normal, marginal and abnormal were 67. 0%, 20. 4% and 12. 6% respectively. The average score of hyperactive attention deficit was 4. 56±2. 32 points, and the proportions of normal, marginal and abnormal people were 69. 4%, 13. 6% and 17. 0% respectively. The average score of prosocial behavior was 6. 38±1. 76 points. The detection rates of normal, marginal and abnormal behaviors were 78. 6%, 16. 1% and 5. 3% respectively. The total score of difficulties was 11. 83 ± 3. 57 points, and the proportions of normal, marginal and abnormal people were 70. 8%, 18. 0% and 11. 2% respectively. The average score of internalized behavior was 5. 17 ± 2. 34 points; The average score of externalized behavior was 6. 66 ± 2. 42 points.

**Table 5 Basic situation of emotional and behavioral problems of preschool children**

### Changes in emotional and behavioral problems after component isochronous substitution

According to the 15-minute isochronic substitution model (Table 6), it was found that after the 15-minute MVPA “one-to-one” isochronic substitution of SP and SB, the internalized behavior score, externalized behavior score and total difficulty score of preschool children were all significantly reduced. When 15 minutes of MVPA and others replaced LPA, only the total difficulty score of preschool children showed a significant downward trend. On the contrary, after replacing MVPA with SP or SB at 15 minutes, the internalized behavior score, externalized behavior score and total difficulty score of preschool children all increased significantly. However, after the 15-minute LPA isochronism replaced the MVPA, in addition to a significant decrease in the total score of difficulty among preschool children, the score of externalized behavior also showed a significant downward trend. Furthermore, after the 15-minute SP isochrone replaced LPA, both the externalized behavior scores and the total difficulty scores of preschool children increased significantly. Conversely, it will be significantly reduced. The mutual substitution among 24-hour activity behaviors has no significant effect on the prosocial behavior scores of preschool children.

**Table 6 Estimated difference in total score of Executive Function for 15-minute isochronous substitution between 24-hour movement behaviors**

Note: * indicates P<0. 05; The data in the table all represent the relationship with the scores of each dimension of the execution function. “-” indicates a negative correlation.

Figure 2 reflects the expected impact of isochronous substitution of different behaviors on the total difficulty score of preschool children. Since the arithmetic mean of MVPA in the research data of this study is 34. 24min, this study takes -30min to 30min of a single activity behavior as the replacement interval and 5min as the unit increment to draw the predicted change trend graph of the influence of equal substitution between ppairs of activity behaviors on the total difficulty score of preschool children. Based on the above figure and the predicted change table, as the time for MVPA to replace SP, SB, and LPA increases, the difficulty score of preschool children shows a downward trend, indicating that emotional and behavioral problems will be improved. When the replacement time was 5 minutes, the total difficulty scores of preschool children decreased by 0. 29, 0. 27, and 0. 22 units respectively. When the replacement time increased from 10 minutes to 30 minutes, the total difficulty scores of preschool children increased by 0. 54-1. 35, 0. 50-1. 22, and 0. 40-0. 91 units respectively. With the extension of the substitution time, the decreasing trend of MVPA replacing other activity behaviors in the total difficulty score of preschool children gradually slows down. As the time for SP, SB and LPA to replace MVPA increases, the difficulty score of preschool children shows an upward trend, indicating that there will be a deepening trend in predicting emotional and behavioral problems. With the extension of the replacement time, the increase rate of the total difficulty score of preschool children gradually accelerates.

**Figure 2.**
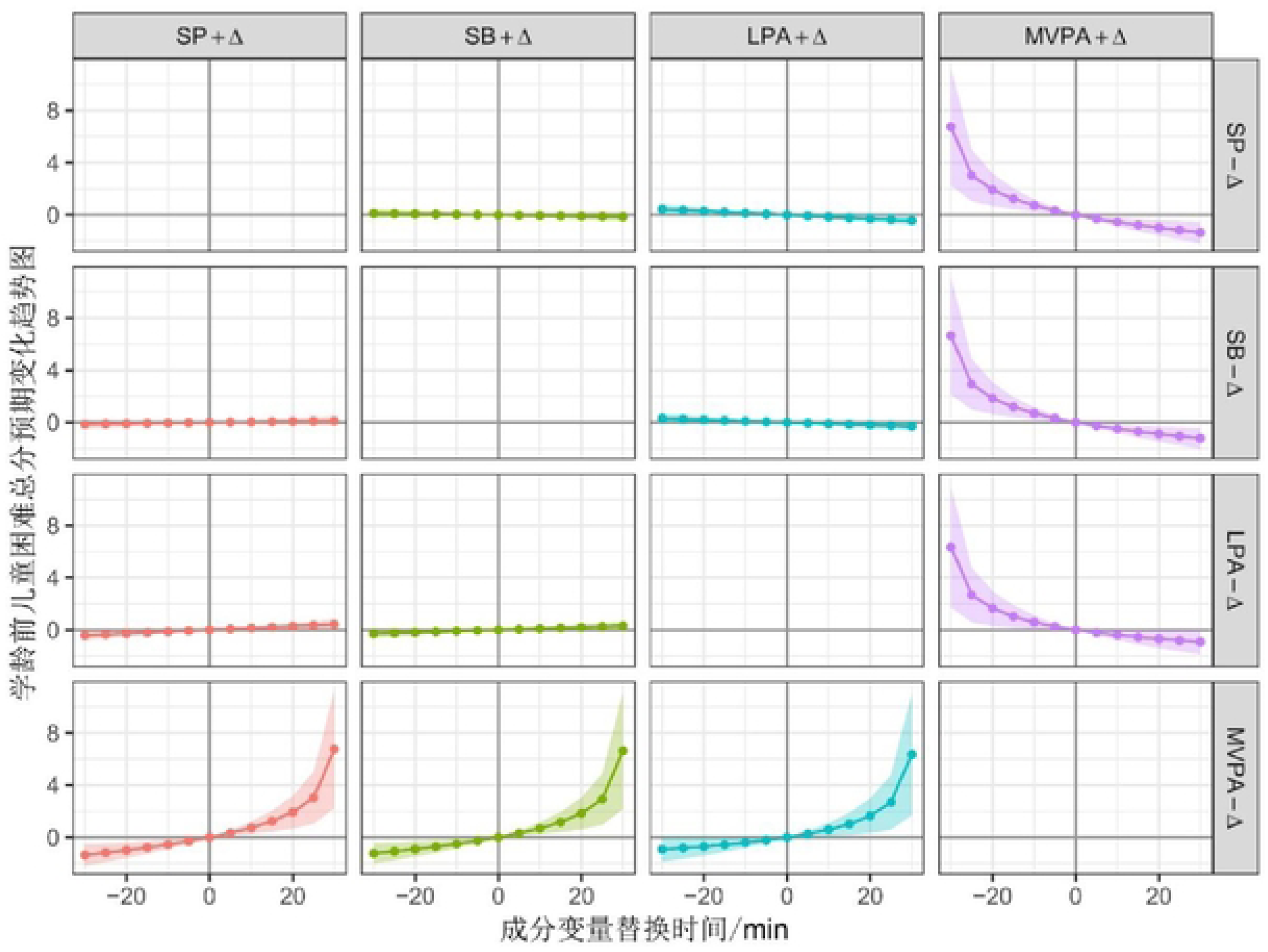
The changing trend of the influence of isochronous substitution on emotional and behavioral problems of preschool children. Note: “+Δ” indicates an increase in the duration of the activity behavior, “-Δ” indicates a decrease in the duration of the activity behavior, and the shaded area represents the 95% confidence interval.

## Discussion

The average daily SP, SB, LPA and MVPA of preschool children in Zhuzhou area investigated in this study were 610. 1min, 438. 3min, 356. 6min and 35. 0min respectively. Compared with the recommended amount in the “24-Hour Activity Guidelines for Early Childhood (0-4 years old) in Canada” issued by Canada in 2017, the MVPA time of preschool children in this study was seriously insufficient. PA for preschool children is mainly carried out in two forms: low-intensity indoor activities and high-intensity outdoor activities [19]. During the investigation period of this study, the weather was bad with continuous rain, and there was also spring flu. Many preschool children had colds and fevers. The weather and disease factors restricted the preschool children in this study from engaging in high-intensity outdoor activities and instead turned to low-intensity indoor activities. In addition, this study found that there were significant gender differences in the use of 24-hour activity and behavior time among preschool children. The MVPA time of male students (40. 9min/d) was significantly higher than that of female students (30. 6min/d), which was consistent with the research results of quan [20] et al. Crespo [21] et al. found that family, society and environment are important factors leading to gender differences in MVPA in young children. Future studies need to fully consider the gender differences of PA in young children. In terms of age, this study found that the MVPA time of preschool children increased with the increase of age. This might be due to the fact that the development level of motor skills of preschool children increases with age. The higher the development level of motor skills, the higher the willingness of preschool children to participate in PA will be, and they will also master some fine motor skills better, and the intensity of PA will also be correspondingly improved [22]. This study also found that the SP time of preschool children decreased with the increase of age, which was consistent with the results of previous studies [23]. The sleep duration of preschool children is negatively correlated with age, which is in line with the physiological development law of young children [24]. It can be seen from the results of the mutation matrix of this study that the 24-hour activity behaviors are interdependent, and there is a possibility of mutual transformation among the various activity behaviors. Among all the behaviors, the pairwise logarithmic ratio of SP and SB is the smallest, indicating that the conversion between these two behaviors is the most likely to occur. The reason for this is that, on the one hand, the health benefits of SP and less SB have been confirmed [8, 9]. Some families, in order to cultivate a healthy lifestyle in preschool children, usually reduce the time spent on screens and other distracting devices, allowing children to go to bed early and get up early. On the other hand, many families, in order to cultivate their children’s cultural literacy, may reduce the night SP time of preschool children and increase their study SB time. Furthermore, the pairwise logarithmic ratios of MVPA to the other three activity behaviors are all relatively large, indicating that the time of MVPA is relatively stable and not prone to transformation. If there are any changes, it is most likely to be converted to LPA. It is not difficult to understand that if one stops to rest immediately after MVPA, there is a high possibility of motor shock. Therefore, after MVPA, some simple LPA should be done before taking a break. This might be the reason why MVPA and LPA are most prone to conversion.

The 15-minute isochronic substitution results show that the mutual substitution between MVPA and other activity behaviors (SP, SB, LP), as well as between SP and LPA, has a significant impact on reducing the scores of internalized behaviors, externalized behaviors, and the total score of difficulty. Moreover, there are certain differences in the predicted change values of emotional and behavioral problems after mutual substitution among different behaviors. This research result reveals that Reasonable allocation of the 24-hour activity and behavior time is of great significance for improving the emotional and behavioral problems of preschool children. It is worth noting that the isochronic substitution results of this study show that 15-minute MVPA or LPA replacing SP can significantly reduce the internalized behavior score, externalized behavior score and total difficulty score of preschool children. However, the average daily SP time of preschool children in this study was 610. 1min/d, which was only 10min more than the recommended SP time in the Canadian guidelines (600-780min). If the 15-min SP was replaced by PA, the SP time of preschool children would not meet the health standard. Zheng [25] et al. Longitudinal studies have found that the duration of sleep at night is negatively correlated with emotional and behavioral problems in preschool children. The mutual substitution between MVPA or LPA and SP requires careful consideration. For preschool children with relatively sufficient sleep time, replacing SP with MVPA or LPA seems to be a good choice for improving emotional and behavioral problems. However, for young children with insufficient SP time, within the limited time of a day, the SP time should not be transferred to PA activities without a lower limit. To avoid the negative impact of severe insufficiency of SP causing emotional and behavioral problems being greater than the positive benefit of PA on emotional and behavioral issues. In addition, MVPA isotime replaces SB and has significant improvement benefits for the internalized behavior score, externalized behavior score and total difficulty score of preschool children. A domestic cohort study found that children who watch videos for too long are more likely to have emotional and behavioral problems in the early stage, and excessive video duration is a risk factor for emotional and behavioral problems in children [26]. Another study found that the duration of video was significantly negatively correlated with the SP time of preschool children [27]. With the extension of video time, the SP time of young children will decrease, thereby indirectly affecting their emotional and behavioral problems. Relevant research has found that the regulation of emotional behavior by PA is mainly related to the transmission of neurotransmitters. For example, during the exercise process of preschool children, the body releases neurotransmitters such as dopamine, norepinephrine and endorphin. These three neurotransmitters are involved in the regulation of emotions and have the effect of alleviating negative emotions. Moreover, the higher the level of PA, the better The more neurotransmitters are released by the body [28, 29]. This study also confirmed this view. The 15-minute mVPA isochronic replacement of LPA has a significant improvement effect on the total difficulty score of preschool children. Based on the significant effects obtained by MVPA replacing other behaviors in the isochronic substitution model, kindergartens should pay attention to the harm caused by the insufficiency of MVPA in preschool children. During their time in the kindergarten, the SP time can be reduced and indoor and outdoor activity courses can be increased to promote the conversion of SB to MVPA time. Meanwhile, gradually increase the intensity of exercise to promote the transition from LPA to MVPA in preschool children, thereby maintaining a better emotional and behavioral level for preschool children.

From the overall perspective of the component isochronic substitution model, the predicted value changes of the total difficulty score by MVPA isochronic substitution of the other three activity behaviors are asymmetric. As the time for MVPA to replace SP, SB and LPA increases, the decline in the total score of difficulty tends to be gentle. However, as MVPA is replaced by SP, SB and LPA, the total score of difficulty for preschool children increases rapidly. This asymmetry is due to the fact that the 24-hour activity behavior data, as component data, conveys relative information rather than absolute information [30]. Specifically, the replacement volume of MVPA at 15 minutes accounted for 42. 9% of the total MVPA of preschool children in this study, while the SP, SB and LPA at 15 minutes accounted for 2. 5%, 3. 4% and 4. 2% respectively. From this, it can be seen that the proportion of the replacement volume of MVPA at 15 minutes is much higher than the ratios of SP, SB and LPA. This leads to the asymmetry after MVPA substitutes for other behaviors. This feature further illustrates the significance of MVPA for emotional and behavioral problems in preschool children. Therefore, educators need to pay close attention to the negative impact of insufficient MVPA in preschool children on their emotional and behavioral problems, gradually mobilize the enthusiasm of preschool children for activities, follow the principle of gradual progress, and gradually transition to the recommended MVPA amount within 60 minutes.

## Research Limitations

Based on the component data analysis method, this study regards the 24-hour activity behavior as a whole and explores the impact of isochrone substitution among different activity behaviors on the emotional and behavioral problems of preschool children, providing specific suggestions for future changes in the direction of 24-hour activity behavior. To ensure the accuracy of the data in this study, an objective measurement method using an accelerometer was adopted. Meanwhile, there were many control variables. However, this study still has the following limitations: (1) The sample size of this study was all collected from the same prefecture-level city, which has regional limitations and cannot fully represent the overall level of preschool children. (2) This study is a cross-sectional one. The data analysis results can only reflect the correlation between variables but not the causal relationship. (3) The ActiGraph wGT3-BT accelerometer is unable to identify the types of sedentary behavior and is unable to explore the influence of different types of sedentary behavior on emotional and behavioral problems. (4) This study, based on the isochronic substitution of component data, explores the theoretical expected impact of isochronic substitution among different activity behaviors on the emotional and behavioral problems of preschool children. More intervention studies should be conducted in the future to investigate the impact of changes in different activity behaviors on emotional and behavioral problems.

## Data Availability

All relevant data are within the manuscript and its Supporting Information files.

## Supporting information

## Acknowledgments

The authors are grateful to the Staff members of Physical Education Department of Wuxi Taihu University

## Author Contributions

**Conceptualization:** Qiong Shen, Aolin Yu, QIANG Lei, CHEN Kai

**Data curation:** Qiong Shen, Aolin Yu

**Formal analysis:** Aolin Yu

**Investigation:** Qiong Shen

**Methodology:** Qiong Shen

**Validation:** Aolin Yu

**Writing – original draft:** Aolin Yu

**Writing – review & editing:**Aolin Yu, Qiong Shen

